# Net Clinical Benefit of Edoxaban for 12 versus 3 Months in Cancer-associated Isolated Distal Deep Vein Thrombosis: ONCO DVT study

**DOI:** 10.1101/2024.02.27.24303473

**Authors:** Yuji Nishimoto, Yugo Yamashita, Takeshi Morimoto, Nao Muraoka, Michihisa Umetsu, Takuma Takada, Yoshito Ogihara, Tatsuya Nishikawa, Nobutaka Ikeda, Kazunori Otsui, Daisuke Sueta, Yukari Tsubata, Masaaki Shoji, Ayumi Shikama, Yutaka Hosoi, Yasuhiro Tanabe, Ryuki Chatani, Kengo Tsukahara, Naohiko Nakanishi, Kitae Kim, Satoshi Ikeda, Yukihito Sato, Tetsuya Watanabe, Takahisa Yamada, Masatake Fukunami, Takeshi Kimura, ONCO DVT Study Investigators

**Affiliations:** Division of Cardiology, Osaka General Medical Center, Osaka, Japan; Department of Cardiology, Hyogo Prefectural Amagasaki General Medical Center, Amagasaki, Japan; Department of Cardiovascular Medicine, Graduate School of Medicine, Kyoto University, Kyoto, Japan; Department of Clinical Epidemiology, Hyogo Medical University, Nishinomiya, Japan; Division of Cardiology, Shizuoka Cancer Center, Shizuoka, Japan; Division of Vascular Surgery, Department of Surgery, Tohoku University Hospital, Sendai, Japan; Department of Cardiology, Tokyo Women’s Medical University, Tokyo, Japan; Department of Cardiology and Nephrology, Mie University Graduate School of Medicine, Tsu, Japan; Department of Onco-Cardiology, Osaka International Cancer Institute, Osaka, Japan; Division of Cardiovascular Medicine, Toho University Ohashi Medical Center, Tokyo, Japan; Department of General Internal Medicine, Kobe University Graduate School of Medicine, Kobe, Japan; Department of Cardiovascular Medicine, Graduate School of Medical Sciences, Kumamoto University, Kumamoto, Japan; Department of Internal Medicine, Division of Medical Oncology and Respiratory Medicine, Shimane University Faculty of Medicine, Izumo, Japan; Department of Cardiovascular Medicine, National Cancer Center Hospital, Tokyo, Japan; Department of Obstetrics and Gynecology, Faculty of Medicine, University of Tsukuba, Tsukuba, Japan; Department of Cardiovascular Surgery, Kyorin University Faculty of Medicine, Tokyo, Japan; Department of Cardiology, St Marianna University School of Medicine, Kawasaki, Japan; Department of Cardiovascular Medicine, Kurashiki Central Hospital, Kurashiki, Japan; Division of Cardiology, Fujisawa City Hospital, Fujisawa, Japan; Department of Cardiovascular Medicine, Graduate School of Medical Science, Kyoto Prefectural University of Medicine, Kyoto, Japan; Department of Cardiovascular Medicine, Kobe City Medical Center General Hospital, Kobe, Japan; Department of Cardiovascular Medicine, Nagasaki University Graduate School of Biomedical Sciences, Nagasaki, Japan; Department of Cardiology, Hirakata Kohsai Hospital, Hirakata, Japan

**Keywords:** venous thrombosis, neoplasms, edoxaban, hemorrhage, recurrence

## Abstract

**Background:** The ONCO DVT (Edoxaban for 12 Months Versus 3 Months in Patients With Cancer With Isolated Distal Deep Vein Thrombosis) study has revealed the superiority of a 12-month versus 3-month edoxaban treatment in terms of fewer thrombotic events for cancer-associated isolated distal deep vein thrombosis; however, concern for increased bleeding with longer anticoagulation remains.

**Methods:** In this post-hoc analysis of the ONCO DVT study, we compared 12-month and 3-month edoxaban treatments in terms of the net adverse clinical events (NACE) as a composite endpoint of the primary endpoint (symptomatic recurrent VTE and VTE-related death at 12 months) and major secondary endpoint (major bleeding at 12-months) of the ONCO DVT study. The net clinical benefit of a 12-month over 3-month treatment was defined as the sum of the differences in the incidence of thrombotic and bleeding events between the 3-month and 12-month treatments. The weight of bleeding events was set at 1.0, and we assessed the changes in the net clinical benefit with weights of bleeding events set at 0.5 and 2.0.

**Results:** With a weight of bleeding events of 1.0, NACE occurred in 30 of 296 patients (10.1%) in the 12-month edoxaban group and in 42 of 305 patients (13.8%) in the 3-month edoxaban group. The net clinical benefit was 3.6% (95% CI, -1.5% to 8.8%). There was a significant treatment-by-subgroup interaction between the thrombocytopenia or cancer metastasis subgroup factors and the effect of the 12-month relative to the 3-month treatment for NACE. As the weights of bleeding events changed from 0.5 to 2.0, the net clinical benefit in the 12-month edoxaban group as compared to the 3-month edoxaban group became attenuated from 4.8% (95% CI, 0.5% to 9.0%) to 0.7% (95% CI, -5.7% to 7.1%).

**Conclusions:** The net clinical benefit of the 12-month over 3-month edoxaban treatment was not significant; however, the 12-month treatment had a numerically lower incidence of NACE than the 3-month treatment.

**Clinical Trial Registration:** ClinicalTrials.gov, NCT03895502.

**Clinical Perspective:** *What is new?:*

- The net clinical benefit of the 12-month over 3-month edoxaban treatment was not significant in terms of the net clinical adverse events (NACE) combined with symptomatic recurrent venous thromboembolism (VTE), VTE-related death, or major bleeding with a weight of bleeding events of 1.0, however, the 12-month group had a numerically lower incidence of the NACE than the 3-month group.
- The net clinical benefit of the 12-month over 3-month edoxaban treatment became attenuated as the weights of the bleeding events increased.

*What are the clinical implications?:*

- The present study revealed that the 12-month edoxaban treatment compared with the 3-month edoxaban treatment was basically favorable in terms of NACE; however, the net clinical benefit of the 12-month edoxaban treatment became attenuated as the weights of the bleeding events increased.
- Further studies should be required to evaluate the case fatality rate of each event and its impact on cancer treatment.

## Introduction

Cancer-associated venous thromboembolism (VTE) is a major concern in the cancer trajectory because it is one of leading causes of death in patients with cancer and also can interrupt or delay cancer treatment.^1,2^ In addition, the incidence of cancer-associated VTE has increased in recent years due to an improved prognosis in patients with cancer, increased use of thrombogenic anticancer-drugs, growing awareness of cancer-associated thrombosis, and more frequent imaging tests for cancer assessments.^1,3–5^ Recent randomized controlled trials comparing direct oral anticoagulants (DOACs) with low-molecular-weight heparin have revealed that DOACs could be a potential treatment option for cancer-associated VTE, which has resulted in a paradigm shift in the treatment strategies.^6–11^ Given the long-term high risk of recurrence in patients with cancer-associated VTE,^12^ the latest guidelines recommend prolonged anticoagulation therapy including DOACs for these patients.^13–16^ However, patients with cancer also have a high risk of bleeding during anticoagulation therapy, leading to a challenge in achieving a risk-benefit balance with anticoagulation therapy.^17^

The recent ONCO-DVT (Edoxaban for 12 Months Versus 3 Months in Patients With Cancer With Isolated Distal Deep Vein Thrombosis) study showed the potential benefit of extended anticoagulation therapy with edoxaban in patients with cancer and isolated distal deep vein thrombosis (DVT), whereas it also suggested a potential signal for an increased risk of bleeding with a longer duration of edoxaban treatment.^18^ It would be clinically relevant to consider the risk-benefit balance of the duration of anticoagulation therapy in cancer-associated VTE. Therefore, we conducted a post-hoc analysis on an exploratory basis to compare the net adverse clinical events (NACE) combining thrombotic and bleeding events between a 12-month and a 3-month edoxaban treatment in patients with cancer-associated isolated distal DVT using the ONCO DVT study database.

## Methods

### Design and ethical statements

The ONCO DVT study was an investigator-initiated, multicenter, open-label, adjudicator-blinded, superiority, randomized controlled trial conducted at 60 institutions in Japan that was designed to compare 12-month and 3-month edoxaban treatment regimens in patients with cancer and isolated distal DVT.^18^ The details of the study organization and participating centers are provided in **Supplementary Appendix 1** and **2**, respectively. The full details of the ONCO DVT study were previously described.^18^ Briefly, patients with active cancer who were newly diagnosed with isolated distal DVT using ultrasonography were randomly assigned, in a 1-to-1 ratio, either to the 12-month or 3-month edoxaban treatment groups. The study was conducted in accordance with the amended Declaration of Helsinki and was approved by the Kyoto University Institutional Review Board and the institutional review boards of all participating centers.

### Study population and treatment

From April 2019 to June 2022, 604 patients were randomized. After excluding 3 patients who withdrew consent during follow-up, 601 patients were included in the present study.

Edoxaban was administered after an appropriate initial treatment after the diagnosis, in accordance with the policies at each institution. No restrictions were set for the policies, providing the treatment did not contravene the exclusion criteria. In the study protocol, edoxaban was recommended to be administered at a reduced dose of 30 mg once daily in patients with a creatinine clearance of 30 to 50 ml per minute or a body weight of 60 kg or less or in those receiving concomitant treatment with potent P-glycoprotein inhibitors.

### Patient characteristics and definitions

Active cancer was defined as cancer meeting one of the following criteria: newly diagnosed within 6 months of randomization; cancer treatment (such as surgery, chemotherapy, or radiotherapy) performed within 6 months of randomization; currently receiving cancer treatment (such as surgery, chemotherapy, or radiotherapy); having a recurrence, local invasion, or distant metastases, or patients with a hematopoietic malignancy who had not achieved a complete remission.^6^ Definitions of the other baseline characteristics are provided in **Supplementary Appendix 3.**

### Clinical outcomes and definitions

In the ONCO DVT study, the primary endpoint was a composite of symptomatic recurrent VTE and VTE-related deaths at 12 months.^18^ The major secondary endpoint was a major bleeding event at 12 months, and other secondary endpoints were symptomatic recurrent VTE events, VTE-related deaths, asymptomatic recurrent VTE events, all clinically relevant bleeding events, and all-cause deaths. Symptomatic recurrent VTE was defined as new or recently worsening pulmonary embolism (PE) or DVT symptoms, new thrombi found on imaging tests, or worsening thrombi compared to the most recent image. Symptomatic VTE recurrences were not determined solely based on the appearance or worsening thrombus images without any new or worsening symptoms. Similarly, if a patient had a thrombus in an index vein with new symptoms, it was not considered as symptomatic recurrent VTE unless a thrombus extension was present. VTE-related deaths were diagnosed at autopsy, following clinically severe PE or death not explained by something other than PE. Major bleeding was defined according to the International Society on Thrombosis and Haemostasis criteria, which was comprised of fatal bleeding, symptomatic bleeding in a critical area or organ, and bleeding reducing the hemoglobin levels by ≥ 2 g/dL or requiring transfusions of ≥ 2 units of whole blood or red cells.^19^ Asymptomatic recurrent VTE was defined as the appearance of new or worsening thrombus images in the pulmonary arteries or deep veins on imaging tests that did not match the definition of a symptomatic VTE recurrence and were not associated with new or worsening symptoms. All clinically relevant bleeding events included major and non-major bleeding events. Clinically relevant non-major bleeding was defined as clinically overt bleeding (including bleeds detected only using imaging) not meeting the criteria for major bleeding, yet that led to one or more of the following: physician-guided medical intervention, hospital admission or further treatment for bleeding, or an in-person medical examination by a physician. The members of an independent clinical outcomes committee who were unaware of the study-group assignments adjudicated all the suspected outcome events and causes of death, as well as the severity of the major bleeding events, with the use of prespecified criteria. The details of the definitions of the other endpoints are described in **Supplementary Appendix 4**.

In the present study, we assessed the risk-benefit balance of a 12-month versus 3-month edoxaban treatment by combining the thrombotic and bleeding events into the NACE. If both thrombotic and bleeding events occurred during the observation period, the first event was considered NACE. We defined NACE-1 as a composite endpoint of the primary endpoint (symptomatic recurrent VTE and VTE-related death at 12 months) and major secondary endpoint (major bleeding at 12 months) of the ONCO DVT study. In addition, to assessing the broader spectrum of thrombotic and bleeding events, we defined NACE-2 as a composite endpoint of symptomatic recurrent VTE, VTE-related death, asymptomatic recurrent VTE, or all clinically relevant bleeding at 12 months.

The net clinical benefit of the 12-month over 3-month edoxaban treatment was defined as the sum of the differences in the incidence of thrombotic and bleeding events between the 3- and 12-month edoxaban treatments.^21^ The net clinical benefit of the 12-month over 3-month edoxaban treatment was calculated as follows: incidence of thrombotic events (3-month edoxaban treatment –12-month edoxaban treatment) + incidence of bleeding events (3-month edoxaban treatment – 12-month edoxaban treatment) × bleeding weight.^21^ The net clinical benefit-1 and net clinical benefit-2 corresponded to NACE-1 and NACE-2, respectively. We set the weight of bleeding events as 1.0 in the base case analyses.

### Statistical analysis

The categorical variables are presented as numbers and percentages and continuous variables as the mean with the standard deviation or median with the interquartile range based on their distributions.

The analysis of the NACE was performed in the intention-to-treat population, which included all the patients who had undergone randomization after excluding those patients who withdrew consent. For patients who did not experience an event, the time to the first event was censored at day 365, or the last day the patient had a complete assessment for the study outcomes, whichever came first. Patients lost to follow-up and patients who died because of reasons other than VTE before the end of the 12-month treatment period and who did not achieve the endpoints, were censored on the last day when the patient had a complete assessment for the study outcomes. We compared the rates of patients with NACE between the 2 treatment groups in the intention-to-treat population and calculated the odds ratios (OR) with the corresponding 95% confidence intervals (CI) using the logistic regression models. The cumulative incidence was estimated using the Kaplan-Meier method, and the differences between the 2 groups were compared using the log-rank test. We also performed subgroup analyses with interaction tests in the prespecified clinically relevant subgroups as well as in the subgroups according to the symptom at diagnosis, which might have had some influence on the thrombotic outcomes. We constructed the same logistic regression models to estimate the P-values for any interaction in the subgroup analyses.

To explore the clinical importance of the bleeding events relative to the thrombotic events, we calculated the net clinical benefit with the weight of bleeding events set at 0.5 and 2.^21^

All reported P values were two-sided and we considered a P <0.05 as statistically significant. All analyses were performed using STATA/SE 17.0 software (StataCorp) and R software version 4.2.2 (R Foundation for Statistical Computing, Vienna, Austria).

## Results

### Patient characteristics

Among the 601 patients included in the present study, there were 296 patients in the 12-month edoxaban group and 305 in the 3-month edoxaban group. The clinical characteristics of the patients at baseline are shown in **Table 1**. The mean age was 70.8 years, 28% of the patients were men, and 20% of the patients had symptoms of DVT at baseline.

**Table 1.**
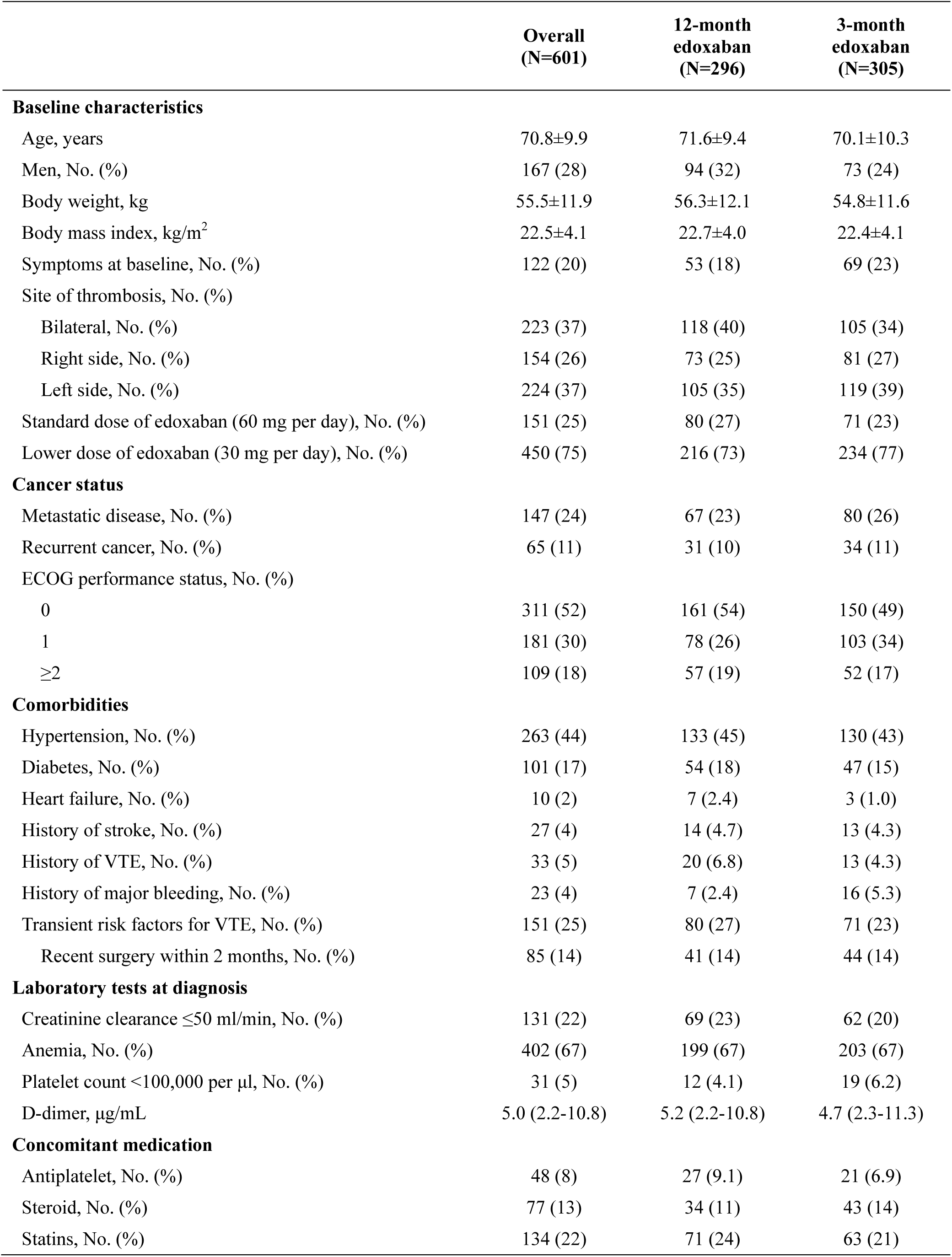
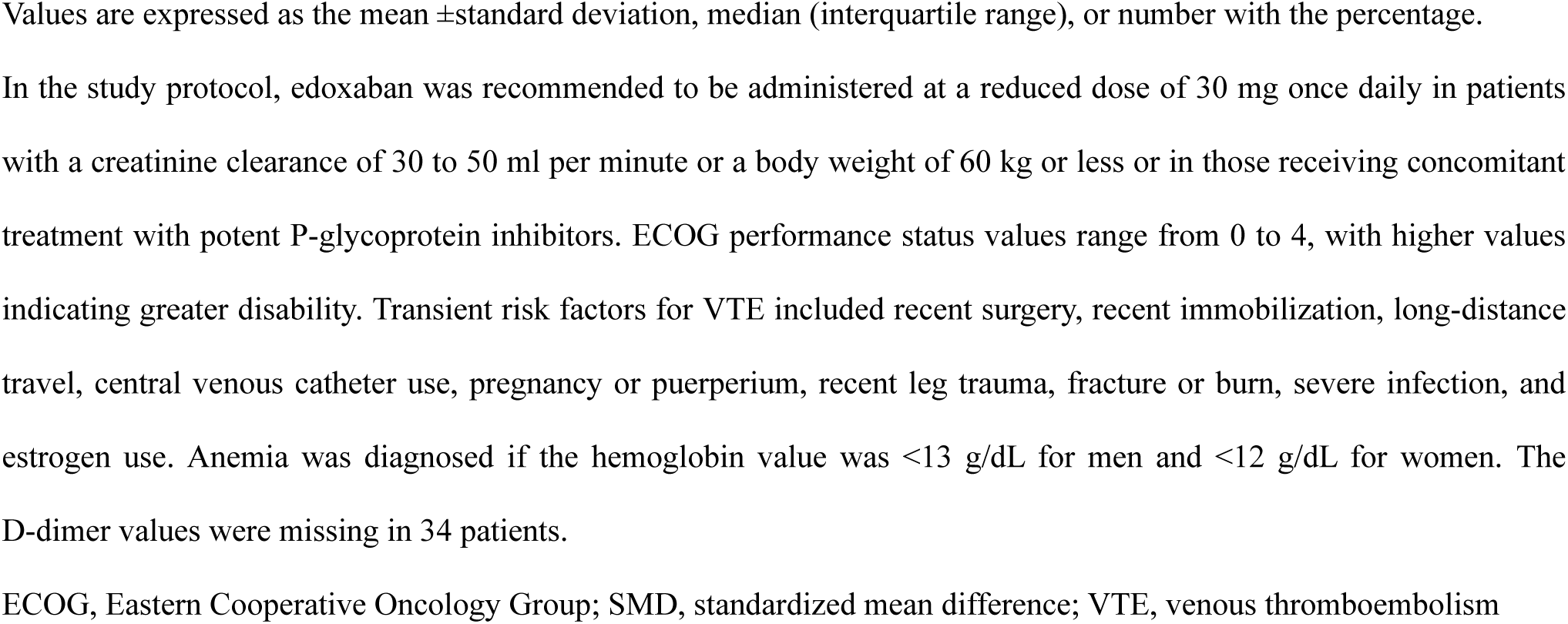
Baseline patient characteristics.

|  | **Overall**<br>**(N=601)** | **12-month**<br>**edoxaban**<br>**(N=296)** | **3-month**<br>**edoxaban**<br>**(N=305)** |
| --- | --- | --- | --- |
| **Baseline characteristics** |  |  |  |
| Age, years | 70.8±9.9 | 71.6±9.4 | 70.1±10.3 |
| Men, No. (%) | 167 (28) | 94 (32) | 73 (24) |
| Body weight, kg | 55.5±11.9 | 56.3±12.1 | 54.8±11.6 |
| Body mass index, kg/m<sup>2</sup> | 22.5±4.1 | 22.7±4.0 | 22.4±4.1 |
| Symptoms at baseline, No. (%) | 122 (20) | 53 (18) | 69 (23) |
| Site of thrombosis, No. (%) |  |  |  |
| Bilateral, No. (%) | 223 (37) | 118 (40) | 105 (34) |
| Right side, No. (%) | 154 (26) | 73 (25) | 81 (27) |
| Left side, No. (%) | 224 (37) | 105 (35) | 119 (39) |
| Standard dose of edoxaban (60 mg per day), No. (%) | 151 (25) | 80 (27) | 71 (23) |
| Lower dose of edoxaban (30 mg per day), No. (%) | 450 (75) | 216 (73) | 234 (77) |
| **Cancer status** |  |  |  |
| Metastatic disease, No. (%) | 147 (24) | 67 (23) | 80 (26) |
| Recurrent cancer, No. (%) | 65 (11) | 31 (10) | 34 (11) |
| ECOG performance status, No. (%) |  |  |  |
| 0 | 311 (52) | 161 (54) | 150 (49) |
| 1 | 181 (30) | 78 (26) | 103 (34) |
| ≥2 | 109 (18) | 57 (19) | 52 (17) |
| **Comorbidities** |  |  |  |
| Hypertension, No. (%) | 263 (44) | 133 (45) | 130 (43) |
| Diabetes, No. (%) | 101 (17) | 54 (18) | 47 (15) |
| Heart failure, No. (%) | 10 (2) | 7 (2.4) | 3 (1.0) |
| History of stroke, No. (%) | 27 (4) | 14 (4.7) | 13 (4.3) |
| History of VTE, No. (%) | 33 (5) | 20 (6.8) | 13 (4.3) |
| History of major bleeding, No. (%) | 23 (4) | 7 (2.4) | 16 (5.3) |
| Transient risk factors for VTE, No. (%) | 151 (25) | 80 (27) | 71 (23) |
| Recent surgery within 2 months, No. (%) | 85 (14) | 41 (14) | 44 (14) |
| **Laboratory tests at diagnosis** |  |  |  |
| Creatinine clearance ≤50 ml/min, No. (%) | 131 (22) | 69 (23) | 62 (20) |
| Anemia, No. (%) | 402 (67) | 199 (67) | 203 (67) |
| Platelet count <100,000 per µl, No. (%) | 31 (5) | 12 (4.1) | 19 (6.2) |
| D-dimer, µg/mL | 5.0 (2.2-10.8) | 5.2 (2.2-10.8) | 4.7 (2.3-11.3) |
| **Concomitant medication** |  |  |  |
| Antiplatelet, No. (%) | 48 (8) | 27 (9.1) | 21 (6.9) |
| Steroid, No. (%) | 77 (13) | 34 (11) | 43 (14) |
| Statins, No. (%) | 134 (22) | 71 (24) | 63 (21) |

Values are expressed as the mean ± standard deviation, median (interquartile range), or number with the percentage.
2 In the study protocol, edoxaban was recommended to be administered at a reduced dose of 30 mg once daily in patients 3 with a creatinine clearance of 30 to 50 ml per minute or a body weight of 60 kg or less or in those receiving concomitant 4 treatment with potent P-glycoprotein inhibitors. ECOG performance status values range from 0 to 4, with higher values 5 indicating greater disability. Transient risk factors for VTE included recent surgery, recent immobilization, long-distance 6 travel, central venous catheter use, pregnancy or puerperium, recent leg trauma, fracture or burn, severe infection, and 7 estrogen use. Anemia was diagnosed if the hemoglobin value was $<13$ g/dL for men and $<12$ g/dL for women. The 8 D-dimer values were missing in 34 patients.
ECOG, Eastern Cooperative Oncology Group; SMD, standardized mean difference; VTE, venous thromboembolism

### Base case analyses

With a weight of bleeding events of 1.0, NACE-1, as a composite of symptomatic recurrent VTE, VTE-related deaths, or major bleeding, occurred in 30 patients (10.1%) in the 12-month edoxaban group and 42 (13.8%) in the 3-month edoxaban group (Log-rank P=0.18; OR 0.71; 95% CI, 0.43 to 1.16) (**Figure 1A and Table 2**). The net clinical benefit-1 of the 12-month over 3-month edoxaban treatment was 3.6% (95% CI, -1.5% to 8.8%).

**Figure 1.**
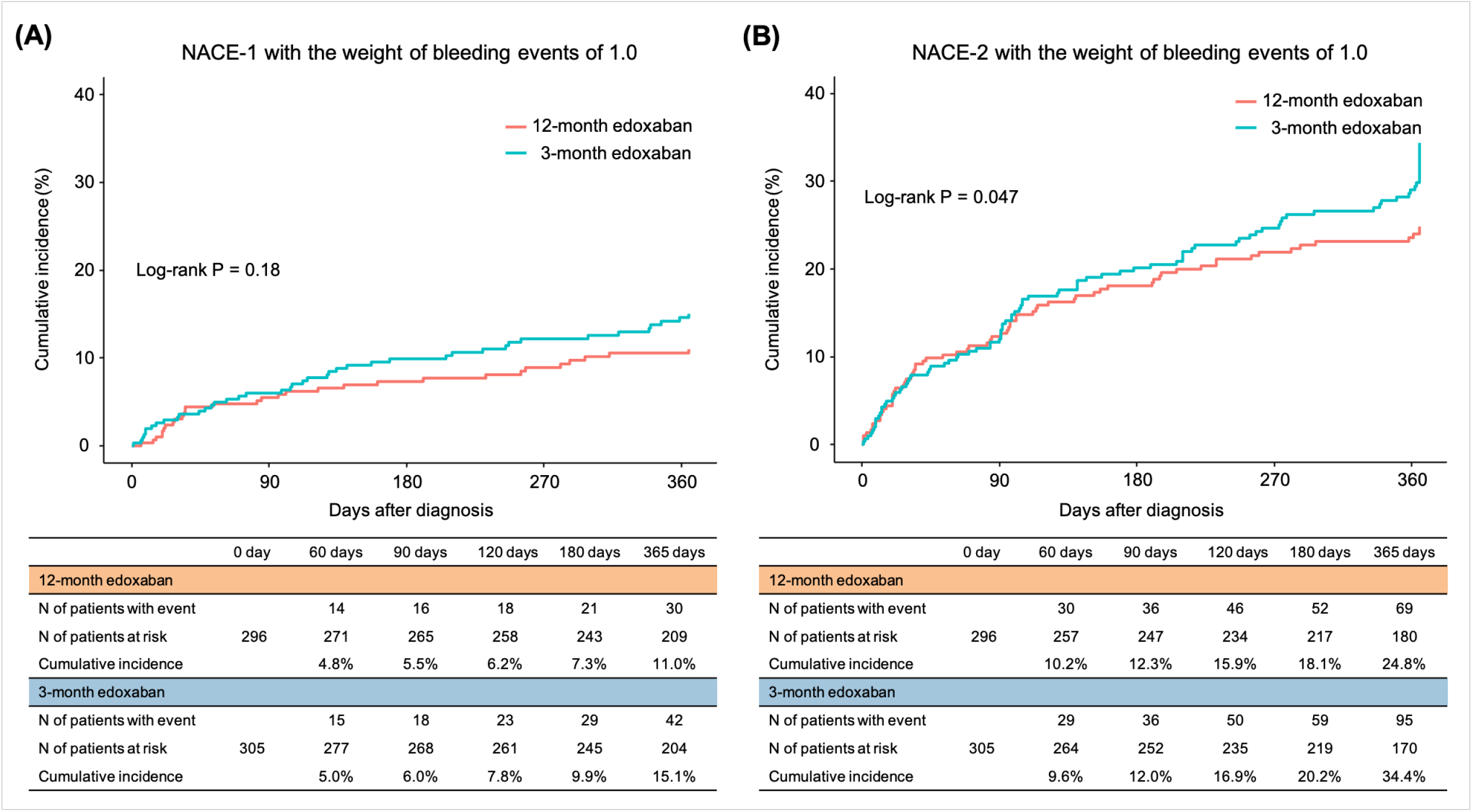
The Kaplan-Meier curves for (A) NACE-1 and (B) NACE-2 comparing the 12-month and 3-month edoxaban treatment groups. Time-to-event curves for (A) NACE-1 (symptomatic recurrent VTE, VTE-related death, or major bleeding) for 12 months and (B) NACE-2 (symptomatic recurrent VTE, VTE-related death, asymptomatic recurrent VTE, or all clinically relevant bleeding) with a weight of bleeding events of 1.0. Asymptomatic recurrent VTE was defined as the appearance of new or worsening thrombus images in the pulmonary arteries and deep veins on imaging tests (ultrasonography of lower limb vein system, computed tomography examination, pulmonary perfusion scintigraphy, pulmonary angiography, and venography) that did not match the definition of a symptomatic VTE recurrence and was not associated with new or worsening symptoms. All clinically relevant bleeding included major bleeding and clinically relevant nonmajor bleeding, which were defined according to the criteria of the International Society on Thrombosis and Hemostasis. NACE, net adverse clinical events; VTE, venous thromboembolism

**Table 2.** Clinical outcomes at 12 months.

|  | 12-month edoxaban<br>(N=296) | 3-month edoxaban<br>(N=305) | Odds ratio<br>(95% CI) |
| --- | --- | --- | --- |
| **Net adverse clinical event-1<sup>†</sup>** | 30 (10.1) | 42 (13.8) | 0.71 (0.43-1.16) |
| Symptomatic VTE or VTE-related death, No. (%) | 3 (1.0) | 22 (7.2) | 0.13 (0.03-0.44) |
| Major bleeding, No. (%)<sup>‡</sup> | 28 (9.5) | 22 (7.2) | 1.34 (0.75-2.41) |
| **Net adverse clinical event-2<sup>†</sup>** | 69 (23.3) | 95 (31.1) | 0.67 (0.47-0.97) |
| Symptomatic VTE or VTE-related death, No. (%) | 3 (1.0) | 22 (7.2) | 0.13 (0.03-0.44) |
| Asymptomatic recurrent VTE, No. (%)<sup>§</sup> | 23 (7.8) | 46 (15.1) | 0.47 (0.28-0.80) |
| All clinically relevant bleeding, No. (%)<sup>‡</sup> | 53 (17.9) | 41 (13.4) | 1.40 (0.90-2.19) |

\*The analyses were performed for the full analysis set based on the intention-to-treat approach, which included all the patients who had undergone randomization after excluding patients who withdrew consent. For patients who did not experience an event, the time to the first event was censored at day 365, or the last day the patient had a complete assessment for study outcomes, whichever came first. We calculated the odds ratios using the logistic regression model along with the corresponding 95% CIs for all clinical endpoints, which have not been adjusted for multiple comparisons.
<sup>†</sup>Net adverse clinical event-1 was a composite of symptomatic recurrent VTE, VTE-related death, or major bleeding with a weight of bleeding events of 1.0. Net adverse clinical event-2 was a composite of symptomatic recurrent VTE, VTE-related death, asymptomatic recurrent VTE<sup>§</sup>, or all clinically relevant bleeding<sup>‡</sup> with a weight of bleeding events of 1.0.
<sup>‡</sup>All clinically relevant bleeding included major bleeding and clinically relevant nonmajor bleeding, which were defined according to the criteria of the International Society on Thrombosis and Hemostasis.
<sup>§</sup>Asymptomatic recurrent VTE was defined as an appearance of new or worsening thrombus images in the pulmonary arteries and deep veins on imaging tests (ultrasonography of lower limb vein system, computed tomography examination, pulmonary perfusion scintigraphy, pulmonary angiography, and venography) that did not match the definition of a symptomatic VTE recurrence and were not associated with new or worsening symptoms.
CI, confidence interval; VTE, venous thromboembolism

NACE-2, as a composite of symptomatic recurrent VTE, VTE-related deaths, asymptomatic recurrent VTE, or all clinically relevant bleeding, occurred in 69 patients (23.3%) in the 12-month edoxaban group and 95 (31.1%) in the 3-month edoxaban group (Log-rank P=0.047; OR 0.67; 95% CI, 0.47 to 0.97) (**Figure 1B and Table 2**). The net clinical benefit-2 of the 12-month over 3-month edoxaban treatment was 7.8% (95% CI, 0.8% to 14.9%). (**Figure 1B**).

### Subgroup analyses

There was no treatment-by-subgroup interaction between the subgroups (pre-specified and presence of symptoms at the time of the diagnosis) and the effect of the 12-month versus 3-month edoxaban treatment for NACE-1, except for those subgroups stratified by the platelet count and cancer metastasis (**Figure 2**). The 12-month edoxaban group had a higher risk of NACE-1 than the 3-month edoxaban group among the patients with thrombocytopenia (platelet count <100,000/μl) (OR 12.9; 95% CI, 1.27 to 131) but a lower risk of NACE-1 among the patients without thrombocytopenia (OR 0.58; 95% CI, 0.34 to 0.98) with a significant interaction (P=0.01). The 12-month edoxaban group had a lower risk of NACE-1 than the 3-month edoxaban group among the patients with cancer metastases (OR 0.34; 95% CI, 0.14 to 0.81) but not among the patients without cancer metastases (OR 1.15; 95% CI, 0.61 to 2.19), and there was a significant interaction (P=0.03) (**Figure 2** and **Supplementary Table S1)**.

**Figure 2.**
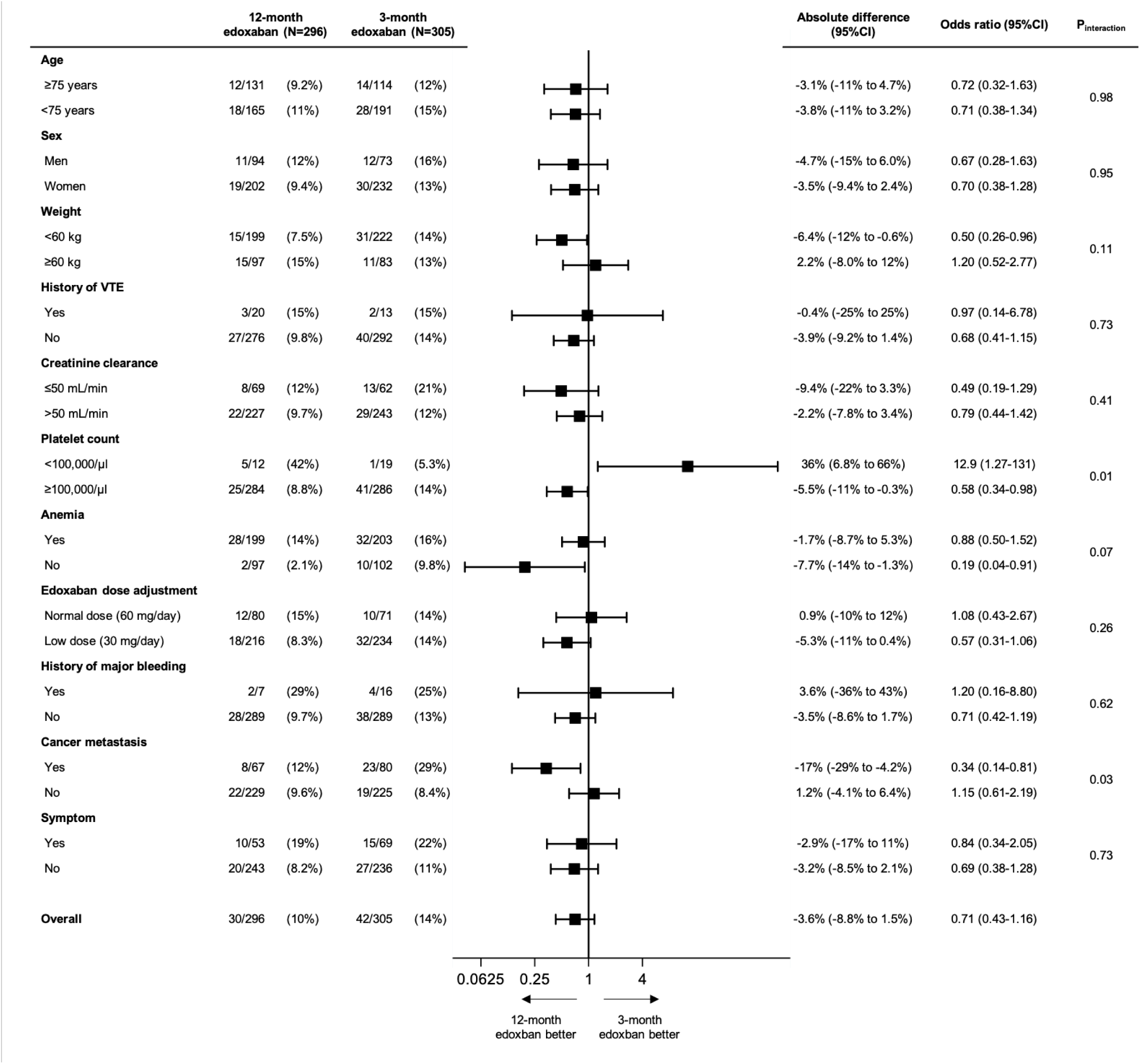
Subgroup analyses for net adverse clinical event-1. Net adverse clinical event-1 was a composite of symptomatic recurrent venous thromboembolism, venous thromboembolism-related death, or major bleeding with a weight of bleeding events of 1.0. The odds ratios with 95% CIs for the net adverse clinical events with the 12-month over 3-month edoxaban treatment are described in the prespecified clinically relevant subgroups as well as in the subgroups according to the symptom at diagnosis. The 95% CIs have not been adjusted for multiple comparisons. CI, confidence interval; VTE, venous thromboembolism

### Effects of the weights of bleeding events on the net clinical benefit

The net clinical benefit according to the weights of bleeding events is shown in **Figure 3**. As the weights of bleeding events increased from 0.5 to 2, the net clinical benefit-1 of the 12-month over 3-month edoxaban treatment became attenuated from 4.8% (95% CI, 0.5% to 9.0%) to 0.7% (95% CI, -5.7% to 7.1%) (**Figure 3A)**. As the weights of bleeding events increased from 0.5 to 2, the net clinical benefit-2 of the 12-month over 3-month edoxaban treatment became attenuated from 10.1% (95% CI, 3.7% to 16.4%) to 3.1% (95% CI, -4.8% to 11.0%) (**Figure 3B**).

**Figure 3.**
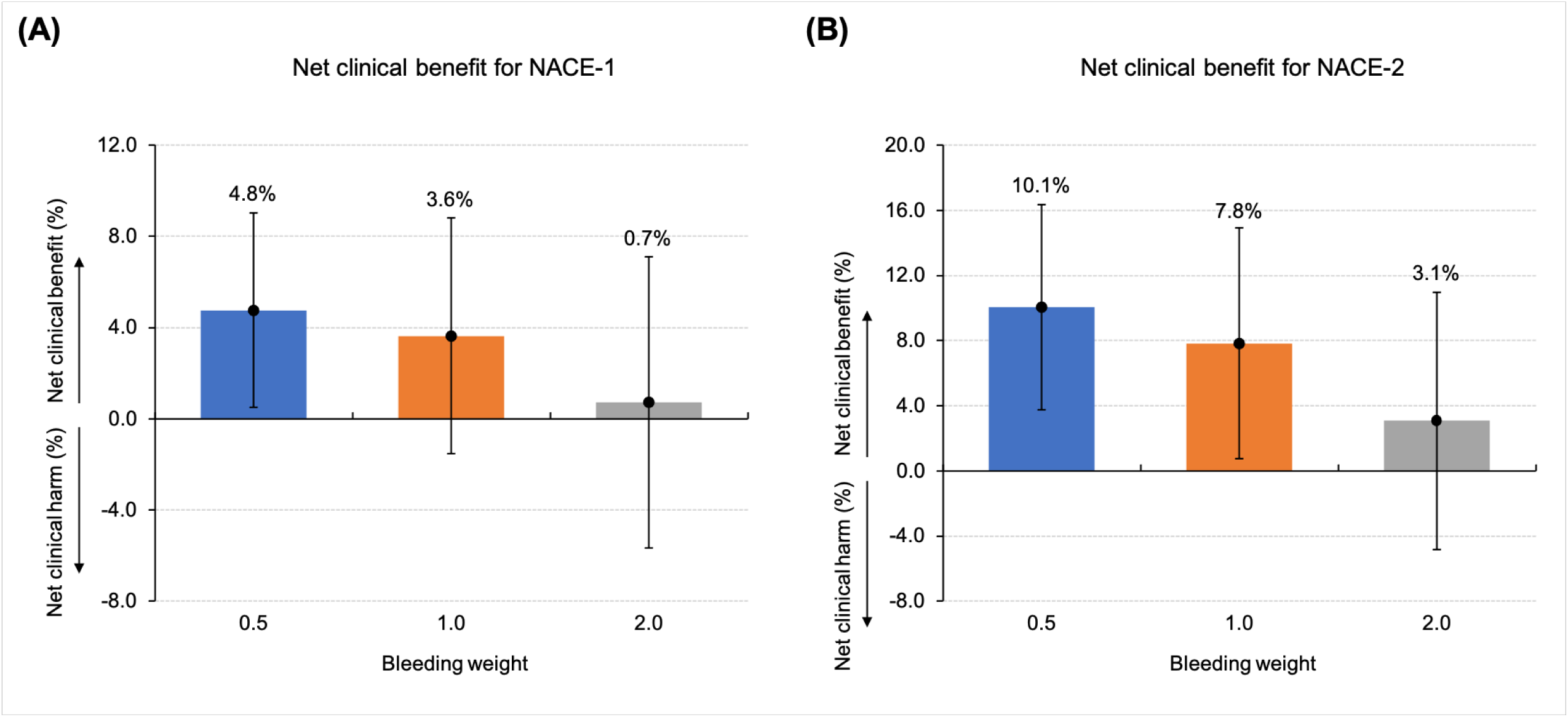
The net clinical benefit of the 12-month over 3-month edoxaban treatment at different bleeding weights: (A) for NACE-1 and (B) for NACE-2. The net clinical benefit of the 12-month over 3-month edoxaban treatment was calculated as follows: incidence of thrombotic events (3-month edoxaban treatment –12-month edoxaban treatment) + incidence of bleeding events (3-month edoxaban treatment – 12-month edoxaban treatment) × bleeding weight. NACE-1 was a composite endpoint of symptomatic recurrent VTE, VTE-related death, or major bleeding at 12 months. NACE-2 was a composite endpoint of symptomatic recurrent VTE, VTE-related death, asymptomatic recurrent VTE, or all clinically relevant bleeding at 12 months. NACE, net adverse clinical events; VTE, venous thromboembolism

## Discussion

The main findings in the present study were as follows: 1) the net clinical benefit of the 12-month over 3-month edoxaban treatment was not significant in terms of NACE-1 combined with symptomatic recurrent VTE, VTE-related death, or major bleeding with a weight of bleeding events of 1.0; however, the 12-month group had a numerically lower incidence of NACE-1 than the 3-month group; 2) in terms of NACE-2 by adding asymptomatic VTE and clinically relevant non-major bleeding to the NACE-1, the net clinical benefit of the 12-month over 3-month edoxaban treatment was significant with a weight of bleeding events of 1.0; 3) the net clinical benefit of the 12-month over 3-month edoxaban treatment became attenuated as the weights of the bleeding events increased; and 4) there was a significant interaction between the subgroups stratified by thrombocytopenia and cancer metastasis and the effect of the 12-month relative to the 3-month edoxaban treatment for NACE.

The ONCO DVT study showed a numerically higher incidence of major bleeding in the 12-month edoxaban treatment (9.5% vs. 7.2%) than in the 3-month edoxaban treatment groups.^18^ Those incidences of major bleeding at 12 months were higher than that in the Hokusai VTE Cancer trial in the edoxaban treatment group (6.9%),^6^ which was partly because the patients in the ONCO DVT study were older and more frequently had renal dysfunction than those in the Hokusai VTE Cancer trial. Because the primary endpoint of the Hokusai VTE Cancer trial was a composite of recurrent VTE or major bleeding, it may be clinically relevant to consider the risk-benefit balance with anticoagulation therapy by assessing the NACE combining thrombotic and bleeding events.^22^ In contrast, there are well-known problems with using composite outcome measures simply as a combination of each event, such as the different clinical importance of each event.^23^ Thus, it would be important to weigh each event in terms of the case fatality rate and impact on daily life.^21–23^ Given the lack of the established weights of events occurring during anticoagulation therapy for VTE, the present study assessed the net clinical benefit using several weights and different composite endpoints on an exploratory basis. The present study revealed that the 12-month edoxaban treatment compared with the 3-month edoxaban treatment was basically favorable in terms of NACE; however, the net clinical benefit of the 12-month edoxaban treatment became attenuated as the weights of the bleeding events increased. Further studies should be required to evaluate the case fatality rate of each event and its impact on cancer treatment.

The present study also showed different effects on the NACE in the subgroups of thrombocytopenia and cancer metastasis. Patients with VTE and thrombocytopenia had a higher risk of major bleeding both in patients receiving warfarin and in those receiving DOACs, especially in the first 3 months of the anticoagulation therapy.^24–26^ Patients with thrombocytopenia in the present study also had a higher risk of major bleeding in the 12-month edoxaban group than in the 3-month edoxaban group, but they had a low risk of thrombotic events in both groups, suggesting the 3-month edoxaban treatment may be beneficial in patients with thrombocytopenia. On the other hand, the previous study showed that the effects of thrombocytopenia on major bleeding may be attenuated in patients with cancer-associated VTE.^26^ Considering the small absolute number of patients with thrombocytopenia in the ONCO DVT study, cautious interpretation would be warranted. For patients with VTE and cancer metastases, the current guidelines recommend prolonged anticoagulation therapy beyond 6 months due to a high risk of VTE recurrence, but supporting evidence is scarce.^2,15^ Those patients also had a high risk of major bleeding during anticoagulation therapy.^27^ The present study showed numerically higher incidences of both thrombotic and bleeding events in patients with cancer metastases than in those without cancer metastases and indicated a potential net clinical benefit of the 12-month over 3-month edoxaban treatment.

The present study had several limitations. First, the open-label design had the potential to introduce bias, including ascertainment bias. However, all clinical endpoints were adjudicated by the members of an independent committee who were blinded to the study group assignments. Second, the present study population included a high proportion of patients with asymptomatic isolated distal DVT. Such a population could be expected to consist of patients with acute DVT, but isolated distal DVT without major symptoms could reflect minor thrombi, which should be interpreted with caution. Third, the adherence to the edoxaban treatment according to the study protocol was relatively low. Particularly, a substantial proportion of the patients in the 12-month edoxaban group discontinued edoxaban prematurely because of bleeding events or cancer progression. Fourth, there could be racial differences in the prevalence of hereditary thrombophilia, a low body weight, and types of cancer, as well as practice differences depending on the regions, such as the ultrasonography methodology. The generalizability of the present results should be carefully assessed. Finally, and most importantly, the present analysis was not a pre-specified analysis, but a post-hoc analysis conducted due to the clinical relevance regarding the numerically higher incidence of major bleeding for the 12-month edoxaban treatment. Thus, the results should be interpreted cautiously as a hypothesis-generating exploratory analysis.

## Conclusions

The net clinical benefit of the 12-month over 3-month edoxaban treatment was not significant; however, the 12-month compared to the 3-month treatment had a numerically lower incidence of NACE.

## Declarations

## Acknowledgments

This study was an investigator-initiated study with financial support from Daiichi Sankyo Co., Ltd. We are grateful to all staff, particularly the Clinical Events Committee, Data Safety Monitoring Committee, and Clinical Research Organization (MID, Inc. and Department of Cardiovascular Medicine, Graduate School of Medicine and Faculty of Medicine Kyoto University) for their thorough work, to all site personnel who helped to enroll participants, and to all health-care professionals who took care of our patients during this study protocol. We also appreciate Mr. John Martin for his grammatical assistance in writing the manuscript. Finally, we thank all of the participants, without whom this research would not have been possible.

## Funding

Funding was provided by Daiichi Sankyo Co., Ltd., which had no role in the study design, data collection, analysis, interpretation, or writing of the report.

## Conflicts of interest

Dr. Nishimoto received lecture fees from Bayer Healthcare, Bristol-Myers Squibb, Pfizer, and Daiichi-Sankyo. Dr. Yamashita received lecture fees from Bayer Healthcare, Bristol-Myers Squibb, Pfizer, and Daiichi-Sankyo, and grant support from Bayer Healthcare and Daiichi-Sankyo. Dr. Morimoto reports lecturer’s fees from AstraZeneca, Bristol-Myers Squibb, Daiichi Sankyo, Japan Lifeline, Kowa, Pfizer and Tsumura; manuscript fees from Bristol-Myers Squibb and Pfizer; advisory board for Novartis and Teijin. Dr. Ogihara received lecture fees from Bayer Healthcare, Bristol-Myers Squibb, Pfizer, and Daiichi-Sankyo, and research funds from Bayer Healthcare and Daiichi-Sankyo. Dr. Dohi received lecture fees from Bayer Healthcare, Bristol-Myers Squibb, Pfizer, and Daiichi-Sankyo, and research funds from Daiichi-Sankyo. Dr. Ikeda N. received lecture fees from Bayer Healthcare, Bristol-Myers Squibb, and Daiichi-Sankyo. Dr. Tsubata received lecture fees from AstraZeneca, Chugai Pharmaceutical, Bristol-Myers Squibb, Kyowa Kirin, Pfizer, Taiho Pharmaceutical, Takeda Pharmaceutical and Daiichi-Sankyo, and grant support from Daiichi-Sankyo, AstraZeneca and OnoPharmaceutical. Dr. Ikeda S. received lecture fees from Bayer Healthcare, Bristol-Myers Squibb and Daiichi-Sankyo. All other authors have reported that they have no relationships relevant to the contents of this paper to disclose.

## Data availability

The data, analytic methods, and study materials will not be made available to other researchers for purposes of reproducing the results or replicating the procedure.

## Ethics approval and consent to participate

The trial was conducted in accordance with the principles of the Declaration of Helsinki and was approved by the Kyoto University Institutional Review Board, along with the institutional review boards of all participating institutions.

## Consent for publication

All patients provided written informed consent.

## Authors’ contributions

**Yuji Nishimoto**: Conceptualization, Methodology, Software, Formal analysis, Investigation, Writing - Original Draft; **Yugo Yamashita**: Conceptualization, Methodology, Software, Formal analysis, Investigation, Writing - Review and Editing; **Takeshi Morimoto**: Conceptualization, Methodology, Writing - Review and Editing; **Takeshi Kimura**: Conceptualization, Methodology, Writing - Review and Editing. All other authors contributed to the Investigation and Supervision. All authors read and approved the final manuscript.

## Notes

### Clinical Trial

ClinicalTrials.gov, NCT03895502.

